# A prediction model based on machine learning for diagnosing the early COVID-19 patients

**DOI:** 10.1101/2020.06.03.20120881

**Authors:** Nan-Nan Sun, Ya Yang, Ling-Ling Tang, Yi-Ning Dai, Hai-Nv Gao, Hong-Ying Pan, Bin Ju

**Affiliations:** Hangzhou Wowjoy Information Technology Co., Ltd, Hangzhou, China; Department of Infectious Diseases, Zhejiang Provincial People’s Hospital, People’s Hospital of Hangzhou Medical College, Hangzhou, China; State Key Laboratory for Diagnosis and Treatment of Infectious Diseases, National Clinical Research Centre for Infectious Diseases, Collaborative Innovation Centre for Diagnosis and Treatment of Infectious Diseases, the First Affiliated Hospital, College of Medicine, Zhejiang University, Hangzhou, Zhejiang Province, 310003, China; Department of Infectious Diseases, ShuLan (Hangzhou) Hospital Affiliated to Zhejiang Shuren University Shulan International Medical College, Hangzhou, China

## Abstract

With the dramatically fast spread of COVID-9, real-time reverse transcription polymerase chain reaction (RT-PCR) test has become the gold standard method for confirmation of COVID-19 infection. However, RT-PCR tests are complicated in operation andIt usually takes 5-6 hours or even longer to get the result. Additionally, due to the low virus loads in early COVID-19 patients, RT-PCR tests display false negative results in a number of cases. Analyzing complex medical datasets based on machine learning provides health care workers excellent opportunities for developing a simple and efficient COVID-19 diagnostic system. This paper aims at extracting risk factors from clinical data of early COVID-19 infected patients and utilizing four types of traditional machine learning approaches including logistic regression(LR), support vector machine(SVM), decision tree(DT), random forest(RF) and a deep learning-based method for diagnosis of early COVID-19. The results show that the LR predictive model presents a higher specificity rate of 0.95, an area under the receiver operating curve (AUC) of 0.971 and an improved sensitivity rate of 0.82, which makes it optimal for the screening of early COVID-19 infection. We also perform the verification for generality of the best model (LR predictive model) among Zhejiang population, and analyze the contribution of the factors to the predictive models. Our manuscript describes and highlights the ability of machine learning methods for improving the accuracy and timeliness of early COVID-19 infection diagnosis. The higher AUC of our LR-base predictive model makes it a more conducive method for assisting COVID-19 diagnosis. The optimal model has been encapsulated as a mobile application (APP) and implemented in some hospitals in Zhejiang Province.

## Introduction

The coronavirus disease 2019 (COVID-19) cases were first reported in Wuhan in December 2019. Soon after, severe acute respiratory syndrome coronavirus 2 (SARS-CoV-2), this new emerging virus has spread rapidly in over 200 countries and areas [1,2]. On March 11, 2020, the World Health Organization (WHO) declared that COVID-19 outbreaks a global pandemic. As of March 25, 2020, COVID-19 has confirmed over 1,946,000 cases and over 126,000 deaths. COVID-19 is a novel pathogen with characteristics of fast transmission and strong infectivity [3,4]. The early symptoms of COVID-19 are similar to other respiratory infectious diseases, which makes it difficult for early differential diagnosis [5-7]. So far, accurate RT-PCR test has been regarded as the gold standard for the diagnosis of COVID-19. However, RT-PCR tests are complicated in operation and it usually takes 5-6 hours or even longer to get the results [8]. Additionally, due to the low virus loads in early infected COVID-19 patients, RT-PCR tests show false negative results in a number of cases [9,10]. It has greatly hindered the prevention and control of the global pandemic. Thus, it is dramatically essential to establish a rapid diagnostic model to screen high-risk patients with COVID-19 infection.

In recent years, machine learning solutions are widely used to predict diagnosis and individual risk factors for diseases, and support clinical decisions [11]. Numerous researchers have adopted different methods in an attempt to improve the precision of data classification in medical field, and a method with superior classification precision would provide better robustness for predicting unknown data [12-14]. Some machine learning methods have achieved remarkable results in medical filed [15,16]. Jagpreet Chhatwal et al. [17] utilized logistic regression to create a breast cancer risk estimation model based on the descriptors of National Mammography Database (NMD) format that can aid in decision-making for early detection of breast cancer. Maggipinto et al. [18] used random forest method to identify the patients who suffer from Alzheimer’s disease based on ADNI datasets, which shows better accuracy and can be used as clinical assistant diagnosis. Recently, the combination of machine learning approaches and epidemic infectious diseases has been emerged extensively. Soyoung Hong et al. [19] used SVM with double class analysis for MERS-COV epidemiological study and discovered the relevance between two sequences of MERS-COV. Wang Jia et al. [20] constructed predictive model with higher accuracy for antigen mutation of influenza virus subtype H1, which used CART decision tree algorithm combined with Amino acid variation sites of viral proteins. The combination of machine learning and medical data has become the main development direction to meet the needs of early diagnosis and prognosis assessment.

In this study, we attempted to identify the best appropriative algorithm for early COVID-19 detection based on clinical big data. We analyzed clinical data of 912 patients who were confirmed as COVID-19 or other respiratory infectious diseases from 18 hospitals in Zhejiang Province, focusing on extraction of risk factors and construction of five types of classification models: SVM, LR, DT, RF as well as deep neural network (DNN). Four epidemiological factors and six clinical manifestations were selected by feature engineering approach as diagnostic models input, and they were much fewer than candidate features of medical records. Essentially, the diagnostic model constructed with fewer meaningful clinical factors is practical for outpatient service. Clinical symptoms, laboratory tests and imaging findings play significant roles in identification of COVID-19 infection [21]. To Evaluate the contributions of clinical symptoms, laboratory tests and imaging information for diagnostic models, we established predictive models based on the data excluding epidemiological information. It was found that the diagnostic models established with clinical symptoms, laboratory tests and imaging information only presented poorer performance. In other words, epidemiological information tremendously affects the performance of COVID-19 predictive models. Briefly, making full use of clinical clinical manifestations and epidemiological characteristics integratedly is essential for constructing the early diagnosis model of COVID-19.

## Materials and methods

### Data construction

The COVID-19 dataset contains clinical information of 914 patients who were confirmed as COVID-19 or other respiratory infectious diseases from 18 hospitals in Zhejiang between Jan 17 and Feb 19, 2020. Considering about the completeness of the clinical information, we firstly screened out the patients with complete clinical records, which results in total number of 912 eligible patients. We then split processed patients into training(80%) and validation(20%) partitions randomly to train our models. Subsequently, we collected 115 clinical dataset from other hospitals in Zhejiang as test partition to verify the universality of implemented models in Zhejiang population.

To obtain the datasets for early stage COVID-19 rapid diagnostic models, all selected patients were categorized into positive or negative cases. The patients who met any one of the following criterias were considered to be positive cases.

- Positive RT-PCR test results in throat swab, sputum, blood samples,
- The genetic sequences detected in the samples are highly homologous to the known SARS-CoV-2.

Positive cases are considered to be the patients confirmed as COVID-19 infection by RT-PCR. Conversely, negative cases are patients excluded as COVID-19 infection by RT-PCR for at least two times. The 912 eligible participants enrolled in this study, include 361 COVID-19 infected patients (positive cases) and 551 COVID-19 non-infected patients (negative cases). Each patient’s clinical record contains 31 factors including gender, age, coexisting diseases, epidemiological informations, laboratory tests, clinical symptoms and imaging findings. Details of these 31 factors and their distribution characteristics on training and validation dataset are shown in table 1.

**Table 1.** The characteristics of positive and negative samples on the training set and validation set.

| *Factors* | Training data set |  | Validation data set |  |
| --- | --- | --- | --- | --- |
|  | positive(n=293) | negative(n=436) | positive(n=68) | negative(n=115) |
| *Gender(Femal)* | 127 | 213 | 31 | 62 |
| *Age(year)* | 47.39±14.38 | 38.53±18.14 | 46.18±14.73 | 35.33±16.85 |
| *Coexistingdiseases* | 102 | 59 | 19 | 17 |
| *Travelorresidencehistoryoverthepast14days* |  |  |  |  |
| *Wuhan* | 97 | 58 | 23 | 12 |
| *NeighboringareasofWuhaninHubeiProvince* | 7 | 53 | 2 | 15 |
| *Otherareaswithpersistentlocaltransmission* |  |  |  |  |
| *orcommunitywithdefinitecases* | 136 | 212 | 26 | 54 |
| *exposuretopatientswithfeverorrespiratory* |  |  |  |  |
| *symptomsoverthepast14days* |  |  |  |  |
| *whohadatravelorresidencehistory* |  |  |  |  |
| *Wuhan* | 77 | 50 | 23 | 14 |
| *NeighboringareasofWuhaninHubeiProvince* | 7 | 18 | 1 | 6 |
| *Otherareaswithpersistentlocaltransmission* |  |  |  |  |
| *orcommunitywithdefinitecases* | 79 | 22 | 20 | 10 |
| *Suspectedpatients* | 4 | 6 | 0 | 5 |
| *Relationshipwithaclusteroutbreak* | 104 | 13 | 29 | 3 |
| *Exposuretowildlife* | 0 | 1 | 1 | 0 |
| *ContactwithpatientsofinfluenzaA* | 1 | 12 | 2 | 3 |
| *ContactwithpatientsofinfluenzaB* | 2 | 11 | 3 | 4 |
| *Bodytemperature* | 37.54±0.852 | 37.48±0.848 | 37.39±0.69 | 37.46±0.73 |
| *Drycough* | 156 | 128 | 32 | 55 |
| *Sputum* | 107 | 104 | 23 | 31 |
| *Fatigue* | 81 | 46 | 17 | 13 |
| *Dyspnea* | 29 | 10 | 4 | 1 |
| *Conjunctivalcongestion* | 1 | 2 | 1 | 2 |
| *Nasalcongestion* | 13 | 37 | 1 | 9 |
| *Diarrheaorbellyache* | 31 | 17 | 6 | 4 |
| *Dizzinessorheadache* | 24 | 38 | 4 | 9 |
| *Nauseaorvomiting* | 10 | 5 | 3 | 2 |
| *Sorethroat* | 15 | 47 | 4 | 14 |
| *Musclesoreness* | 16 | 3 | 1 | 0 |
| *Whitebloodcellcount($\times 10^9$)* | 5.47±2.63 | 7.36±3.04 | 5.10±1.88 | 7.08±2.89 |
| *Lymphocytecount($\times 10^9$)* | 1.22±0.88 | 1.68±0.86 | 1.16±0.54 | 1.81±0.78 |
| *Neutrophilcellcount($\times 10^9$)* | 4.01±4.67 | 5.12±3.47 | 3.47±1.68 | 4.58±2.40 |
| *C – reactiveproteinlevel(mg/L)* | 21.77±27.79 | 18.72±35.26 | 17.33±20.97 | 17.15±34.93 |
| *ImagingchangesofChestX – rayorCT* |  |  |  |  |
| *Normal* | 10 | 194 | 6 | 48 |
| *Unilaterallocalpatchyshadowingl* | 94 | 104 | 13 | 28 |
| *Bilateralmultiplegroundglassopacityl* | 94 | 77 | 24 | 20 |
| *Bilateralwithpulmonaryconsolidation* | 84 | 24 | 25 | 6 |
| *Otherimagingalterations* | 11 | 37 | 0 | 13 |

#### Feature Selection

Feature selection is used to select effective factors from numerous features to reduce the feature space dimension and classification error rate. We leveraged embedded feature engineering approach based on logistic regression algorithm to select COVID-19 risk factors from the 31 factors mentioned above. Finally, 10 factors were chosen for the early COVID-19 prediction task by setting the threshold as 0.85. The final selected factors include four epidemiological features(relationship with a cluster outbreak, travel or residence history over the past 14 days in Wuhan, exposure to patients with fever or respiratory symptoms over the past 14 days who had a travel or residence history in Wuhan, exposure to patients with fever or respiratory symptoms over the past 14 days who had a travel or residence history in other areas with persistent local transmission, or community with definite cases) and six clinical manifestations (muscle soreness, dyspnea, fatigue, lymphocyte count (×10^9^/*L*), white blood cell count (×10^9^/*L*) and imaging changes of Chest X-ray or CT). In practice, the diagnostic model constructed with fewer incoherent factors is beneficial and practical for outpatient service. Details of selected risk factors and their related coefficients are shown in table 2. The importance of the factors relies on absolute value of the coefficients. Table 2 suggests that imaging changes of Chest X-ray or CT is more vital than others. Table 2 also shows that the tolerance of these 10 factors is more than 0.1 and variance inflation factor of them is less than 10, which indicated that there was no collinearity among selected factors.

**Table 2.**
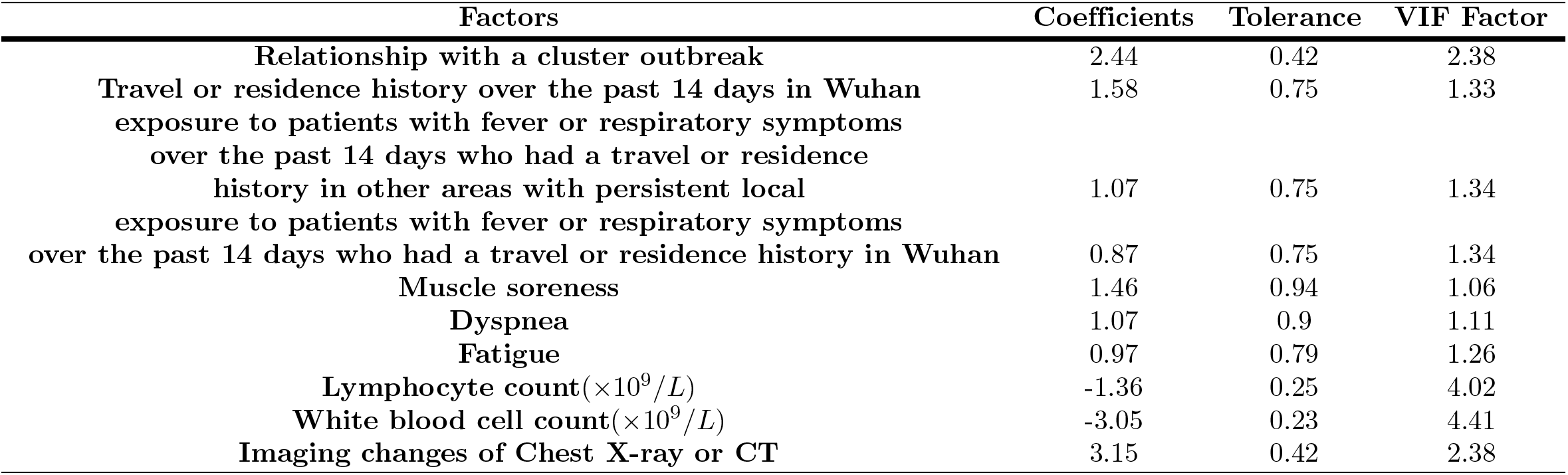
The coefficients, tolerance and variance inflation factor of factors selected by feature engineering.

### Methodology

#### Machine learning models

In this study, we conduct four conventional types of machine learning algorithms and a deep learning solution to establish the early stage COVID-19 rapid diagnostic models. We implement LR model with L2 regularization penalty, and train other three models including SVM with kernel of *rbf*, ID3 DT and FR. The FR model is constructed by 50 decision trees with information gain algorithm. This study used deep learning-base method, namely DNN, which is a four-layer network with the hidden dimension of 64,32 16 and 20 respectively. A Softmax layer is added at the top of the network to output the probability of a patient infected with COVID-19.

We evaluat the performance of the early stage COVID-19 diagnostic models at the 20% validation using familiar assessment strategies, which include measuring accuracy and the AUC generated by plotting sensitivity vs 1 – specificity. Classification accuracy is obtained via an optimum cut-off point. AUC measures the overall performance of the recall concerning different false positive rate, which exhibits robustness for performance assessment of predictive models [22]. Models with higher AUC will show more powerful identified and diagnostic capacities to assist health care workers. High-sensitivity (or recall rate of positive cases) and high-specificity (or recall rate of negative cases) play a vital role in screening the infectious patients [23]. Essentially, a model with high sensitivity can correctly identify patients infected with COVID-19 for timely treatment, while a model with high specificity can excellently screen non-infective patients, thereby effectively avoid cross infection.

## Results

The experiments we conduct to evaluate the performance of the five types of predictive models are illustrated in this section. We evaluate the predictive models on validation set and compare the results of validation to obtain the best solution for identifying early COVID-19 infection. Ultimately, we test the best model based on test dataset to obtain general diagnostic model for Zhejiang population

We implement multiple model structures as our constructed models and deploy different combinations of feature inputs. Table 4 summarizes the performance of conventional solutions and deep learning-based methods. Table 3 part (a) reveals the performances of predictive models constructed based on the raw dataset including 31 factors(table 1), and part (b) exhibits the performances of models established with ten factors selected by using feature selection approach. Feature selection is intended for data dimensionality reduction [24]. In practice, the diagnostic models constructed with less meaningful clinical factors are more practical for outpatient services. The results inTable 3 demonstrate that the predictive models of part (b) perform slightly better than that of part (a) in terms of AUC. The sensitivity, specificity, as well as accuracy of these predictive models of part (b) are relatively approximate to those of part(a). Thus, feature selection partly improves the performances of COVID-19 diagnostic models, and the ROC curve of some selected high performing machine learning models are shown in Fig 1. Table 4 part (b) shows that LR combined with feature selection outperforms other four methods by reaching an AUC of 0.971, high-specificity of 0.95 and accuracy of 0.90 respectively. These results suggest that the combination of LR and feature selection approach presents the best AUC and specificity among five categories of classification methods. Higher specificity of model will facilitate the elimination of infected diseases such as COVID-19 infection. In addition, according to the clinical experience of experts, the AUC (0.86) calculated by the diagnostic scale is compared, and the LR diagnostic model shows better performance. Therefore, LR can be selected as the optimum classification model for the early-stage COVID-19 rapid screening.

**Table 3.** The performance comparison of various machine learning models on validation set with different sets of features.

| Model | Sensitivity | Specificity | Acc | AUC |
| --- | --- | --- | --- | --- |
| (a) models constructed on raw dataset |  |  |  |  |
| LR | 0.84 | 0.95 | 0.91 | 0.967 |
| SVM | 0.74 | 0.87 | 0.82 | 0.916 |
| DT | 0.81 | 0.89 | 0.86 | 0.851 |
| FR | 0.82 | 0.90 | 0.87 | 0.957 |
| DNN | 0.84 | 0.88 | 0.86 | 0.946 |
| (b) models constructed combination with feature selection |  |  |  |  |
| LR+FS | 0.82 | 0.95 | 0.90 | 0.971 |
| SVM+FS | 0.82 | 0.94 | 0.90 | 0.952 |
| DT+FS | 0.85 | 0.90 | 0.88 | 0.884 |
| FR+FS | 0.85 | 0.88 | 0.87 | 0.957 |
| DNN+FS | 0.87 | 0.89 | 0.86 | 0.953 |
| (c) models constructed on the dataset excluding epidemiological information |  |  |  |  |
| LR-exclude epidemiology | 0.76 | 0.85 | 0.82 | 0.872 |
| SVM- exclude epidemiology | 0.71 | 0.87 | 0.81 | 0.871 |
| DT-exclude epidemiology | 0.71 | 0.83 | 0.79 | 0.761 |
| FR-exclude epidemiology | 0.69 | 0.87 | 0.80 | 0.875 |
| DNN-exclude epidemiology | 0.71 | 0.89 | 0.82 | 0.854 |
| (d) models combination with feature selection |  |  |  |  |
| LR+FS-exclude epidemiology | 0.76 | 0.87 | 0.83 | 0.871 |
| SVM+FS-exclude epidemiology | 0.81 | 0.87 | 0.85 | 0.889 |
| DT+FS-exclude epidemiology | 0.71 | 0.88 | 0.81 | 0.815 |
| FR+FS-exclude epidemiology | 0.71 | 0.86 | 0.80 | 0.848 |
| DNN+FS-exclude epidemiology | 0.76 | 0.84 | 0.81 | 0.864 |

Table notes Acc: Accuracy. FS: feature selection. exclude exclude epidemiology: models were established on the dataset excluding epidemiology information.

**Table 4.** The Sensitivity, Specificity, Accuracy, and AUC for logistic regression on test dataset.

| model | Sensitivity | Specificity | Accuracy | AUC |
| --- | --- | --- | --- | --- |
| LR+FS | 0.87 | 0.95 | 0.91 | 0.95 |

Under the background of COVID-19 pandemic, clinical symptoms, laboratory tests and imaging findings are vital clinical criterion for the diagnosis of COVID-19 infection. In order to verify the contribution of above-mentioned three indicators to the COVID-19 diagnostic models, we establish predictive models based on the dataset excluding epidemiological information. The performances of various predictive models are shown in table 3 (c) and (d). Results in part (a) and part (b) illustrates that epidemiological information is beneficial for early COVID-19 rapid diagnostic models construction. In the absence of epidemiological information, the sensitivity, specificity and accuracy of the predictive models (part (c)) exhibits sharp reduction compared with the models shown in part (a). In addition, the five types of machine learning approaches combining with feature selection is constructed based on the dataset excluding epidemiological information, as is shown in Table 4 part (d). Compared with part (c), AUC of part (d) is slightly improved. While due to the absence of epidemiological information, part (c) and part (d) show poorer performances compared with part (a) and part (b). In brief, it indicates laterally that epidemiological information is essential for constructing the early COVID-19 diagnostic models in Zhejiang population.

The above results clearly illustrates that the combination of traditional logistic regression method and feature selection has a great probability to predict early COVID-19 infection. And construction of highly precious diagnostic model relies on integrating and taking the most advantages of clinical symptoms, laboratory tests, imaging findings as well as epidemiological information.

Moreover, LR algorithm is proved as the most ideal method among the five classification solutions for the early COVID-19 rapid screening. The experiments performed in this study used test dataset for verifying generality of the optimum diagnosis model. As is shown in Table 5, the sensitivity, specificity, accuracy and AUC of the LR+ FS model on test dataset are 0.87, 0.95, 0.91 and 0.95, respectively. These results show that the predictive model constructed by combination of logistic regression and feature selection as early COVID-19 rapid diagnostic tool is universally applicable in Zhejiang Province.

## Discussion

Under the background of COVID-19 pandemic, the early prevention and control of COVID-19 still face severe challenges. According to the reports, the most common early symptoms of COVID-19 are fever, cough, fatigue, and myalgia, followed by diarrhea, nausea, headache and sore throat [25,26]. As the disease goes on, some infected patients, especially those with low immune functions, gradually become dyspnea [21,27]. Additionally, complications such as acute arrhythmia and shock, respiratory distress syndrome (ARDS), are probably related to a poor prognosis [28,29]. Thus, early prediction of suspected patients and early aggressive treatment of confirmed patients are the key to reduce cross infection and mortality. CT scan has become the main auxiliary tool for screening of COVID-19 cases. However, CT scan can not be used to identify specific viral infections [30]. Moreover, some COVID-19 patients can also present with normal pulmonary imaging in early stage [31]. Clinical symptoms and laboratory tests are sometimes non-specific for early COVID-19 infection [21,32]. At present, RT-PCR is still the accepted detection method for the diagnosis of COVID-19 infection. While the time consuming and instability of test results are still the most struggling problems [8]. Therefore, to improve the timeliness for the early COVID-19 infection diagnosis, it is essential to develop a decision-making tool to assist early diagnosis of COVID-19 patients in fever clinics.

Current studies which analyze symptoms and laboratory examination results of COVID-19 patients mainly focus on predicting mortality risk and progression of the disease [33]. Only few studies aims at COVID-19 early diagnosis. At present, Zirui Meng et al. [34] selected nine representative variables(including age, Activated Partial Thromboplastin Time, Red Blood Cell Distribution Width-SD, Uric Acid, Triglyceride, Serum Potassium, Albumin/globulin, 3-Hydroxybutyrate, Serum Calcium) and constructed an optimized diagnostic model through Lasso regression screening and Multivariate logistic regression based on 431 samples. The AUC of their early COVID-19 screening model in the testing set and independent validation cohort were 0.890 and 0.872. Cong Feng et al. [35] used logistic regression with Lasso regression for features selection and screening model development based on clinical data of 132 recruited patients. The final chosen features include 1 demographic variable (age); 4 variables of vital signs (e.g., Temperature (TEM), Heart rate (HR), etc.); 5 variables of blood routine values (e.g., Platelet count (PLT), Monocyte ratio (MONO%), Eosinophil count (EO#), etc.); 7 variables of clinical signs and symptoms (e.g., Fever, Fever classification, Shiver, etc.); and 1 infection-related biomarker (Interleukin-6 (IL-6)). The performance of their model constructed based on the final selected features in held-out testing set and validation cohort resulted in AUCs of 0.841 and 0.938, and specificity of 0.727 and 0.778. In our study, we selected four epidemiological features and six clinical manifestations from the raw dataset including 31 factors, further developed multiple models with various machine learning algorithms and screened an optimum early COVID-19 diagnostic model with an AUC of 0.971. We tested the best model based on LR on the external test data set, and its AUC and specificity were 0.950 and 0.95, respectively. Compared to previous studies, we screened out fewer risk factors based on a larger clinical data set, and the early COVID-19 diagnostic model we established has better performance and is more suitable for clinical assisted diagnosis. Moreover, our study is based on a large clinical data set, including a total of 912 patients who were confirmed to have early COVID-19 infection or other respiratory infectious diseases, which may contribute to mining more potential clinical information and improve generalization ability of diagnostic models. Considering the indisputable role epidemiological features play in the diagnosis of infectious diseases in clinic [36], we specifically studied the role of epidemiological information in diagnostic models. We found that the lack of epidemiological information greatly affected the accuracy, specificity and sensitivity of the model. It means that epidemiological information is vital for building an accurate COVID-19 diagnostic tool, and makes the utility and reliability of the previously reported diagnostic models questioned.

Nevertheless, this study still has several limitations. First of all, the recruited participants are limited to Zhejiang Province, which causes certain regional restrictions in the application of the predictive models. Further extremely concerning about the epidemiological characteristics and nationwide studies are needed to access the generality of the suggested model. Secondly, there is a lack of information on the progression and prognosis of COVID-19 as well as asymptomatic infection cases. Finally, more information of infections should be recruited to improve the accurate of screening model.

## Conclusion

In our study, ten representative factors with significant identification value were selected and constructed diagnostic models. The model established an algorithm based on logistic regression can be used as a simple, fast, and effective tool for diagnosing the early COVID-19 infection with significant clinical value.

## Data Availability

all data are fully available without restriction

## Supporting information

**S1 Fig.**
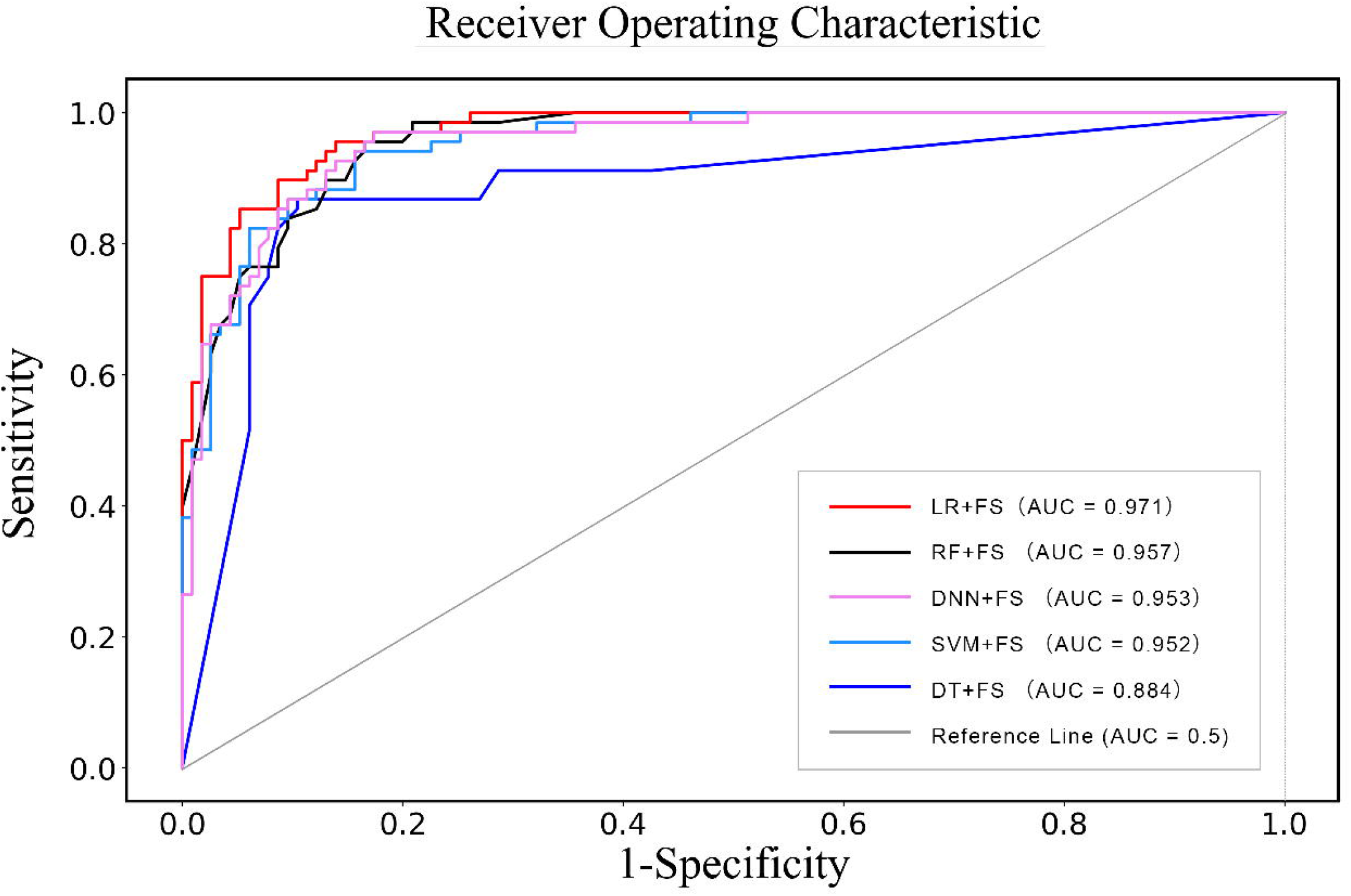
ROC curve. ROC curve of chosen high performing machine learning models.

## Acknowledgments

We thank all the persons who has been fighting in this outbreak. This work was supported by the grant from the emergency project of key research and development plan in Zhejiang Province (2020C03123-2) and National Science and Technology Major Project (2017ZX10204401).

## Notes

### Competing Interest Statement

The authors have declared no competing interest.

### Funding Statement

2020C03123-2 the emergency project of key research and development plan in Zhejiang Province
2017ZX10204401 National Science and Technology Major 253 Project

### Author Declarations

the Ethics Committee of Zhejiang Provincial People's Hospital

## References

1. Z W, JM M. T Characteristics of and Important Lessons From the Coronavirus Disease 2019 (COVID-19) Outbreak in China: Summary of a Report of 72 314 Cases From the Chinese Center for Disease Control and Prevention. JAMA. 2020.

2. Xu Z, Shi L, Wang, Y., et al. T Pathological findings of COVID-19 associated with acute respiratory distress syndrome. The Lancet Respiratory medicine 2020.

3. Chan JF, Yuan S, Kok K. T A familial cluster of pneumonia associated with the 2019 novel coronavirus indicating person-to-person transmission: a study of a family cluster. The Lancet. 2020 2020-01-01;395(10223):514-23.

4. Muhammad A S, Suliman K, Abeer, K., et al. T COVID-19 infection: Origin, transmission, and characteristics of human coronaviruses. Journal of Advanced Research. 2020-03;005(24):91-98.

5. Peeri N C, Nistha S, Siddikur R M, et al. T The SARS, MERS and novel coronavirus (COVID-19) epidemics, the newest and biggest global health threats: what lessons have we learned?. International Journal of Epidemiology. 2020-02-22; https://doi.org/10.1093/ije/dyaa033

6. Deng Congmei. T Causes and Preventive measures of respiratory diseases in autumn and winter. Animal health care in China. 2017;019(001):14.

7. Chavez S, Long B, Koyfman A, et al. T Coronavirus Disease (COVID-19): A primer for emergency physicians. American Journal of Emergency Medicine. 2020.

8. Wang HY. T Nucleic Acid Detection Technology and Enzyme-linked Immune Detection Technology in the Initial Application of Comparative Analysis of Blood Screening. Chinese Journal of Blood Transfusion. 2007;20(1):43–5.

9. Xie XZ, Zhong Z, Zhao, W., et al. T Chest CT for Typical 2019-nCoV Pneumonia: Relationship to Negative RT-PCR Testing. Radiology. 2020;200343.

10. Wu J, Liu J, Li, S., et al. T Detection and analysis of nucleic acid in various biological samples of COVID-19 patients. 2020.

11. Gupta D, Khare S, Aggarwal A. T A method to predict diagnostic codes for chronic diseases using machine learning techniques[C]// 2016 International Conference on Computing. Communication and Automation (ICCCA). IEEE, 2016.

12. Chao CM, Yu YW, Cheng, BW., et al. T Construction the Model on the Breast Cancer Survival Analysis Use Support Vector Machine, Logistic Regression and Decision Tree. MED SYST. 2014;38(10).

13. Yeom S, Giacomelli I, Menaged A, et al. T Overfitting, robustness, and malicious algorithms: A study of potential causes of privacy risk in machine learning. Journal of Computer Security, 2019(3):1–36.

14. Oliveira Bárbara, Daniela G, O’Halloran Martin, et al. T Diagnosing Breast Cancer with Microwave Technology: remaining challenges and potential solutions with machine learning. Diagnostics, 2018, 8(2):36

15. Luo ST, Cheng B. T Diagnosing Breast Masses in Digital Mammography Using Feature Selection and Ensemble Methods. J MED SYST. 2012;36(2):569–77.

16. Fan CY, Chang P, Lin, J., et al. T A hybrid model combining case-based reasoning and fuzzy decision tree for medical data classification. APPL SOFT COMPUT. 2011;11(1):632–44.

17. Chhatwal J, Alagoz OM, Kahn-CE J, et al. T A logistic regression model based on the national mammography database format to aid breast cancer diagnosis. Ajr American Journal of Roentgenology. 2009;192(4):1117–27.

18. Maggipinto T, Bellotti R, Amoroso, N., et al. T DTI measurements for Alzheimer’s classification. Physics in Medicine and Biology. 2017;62(6):2361–75.

19. Hong S, Choi S, Kim D, et al. T Epidemiological analysis of MERS-CoV using NN and SVM in respect to applicability of AI in multiple classes. 2017 19th International Conference on Advanced Communication Technology (ICACT). IEEE. 2017;APPL SOFT COMPUT. 2011;11(1):632–44.

20. Wang J, Wu YL. T Prediction of antigenic variation of influenza virus subtype H1 based on machine learning. Information Communication; 2018;000(009):63–64.

21. Chen N, Zhou M, Dong, X., et al. T Epidemiological and clinical characteristics of 99 cases of 2019 novel coronavirus pneumonia in Wuhan, China: a descriptive study. The Lancet. 2020 2020–01-01;395(10223):507-13.

22. Qin F, Luo H, Cheng, ZK., et al. T Evaluating performance of multiple Bayes classifier based on AUC method. Computer Engineering and Design. 2007;028(24):5919–20.

23. Yu-Wei Lin, Yuqian Zhou, Faraz Faghri, et al. T Analysis and prediction of unplanned intensive care unit readmission using recurrent neural networks with long short-term memory. PLoS ONE. 2018;14(7): e0218942.

24. YIsabelle Guyon, Masoud Nikravesh, Steve Gunn, et al. M Feature Extraction. Springer Berlin Heidelberg, 2006.

25. Han C, Duan C, Zhang, S., et al. T Digestive Symptoms in COVID-19 Patients With Mild Disease Severity: Clinical Presentation, Stool Viral RNA Testing, and Outcomes. Am. J. Gastroenterol. 2020-04-15.

26. Yang X, Zhao J, Yan, Q., et al. T A case of COVID-19 patient with the diarrhea as initial symptom and literature review. Clin Res Hepatol Gastroenterol. 2020 Apr 15.

27. Xu, X.W., et al. T Clinical findings in a group of patients infected with the 2019 novel coronavirus (SARS-Cov-2) outside of Wuhan, China: retrospective case series. BMJ. 2020 2020-02-27:m792.

28. Koff WC, Williams MA. T Covid-19 and Immunity in Aging Populations - A New Research Agenda. N. Engl. J. Med. 2020 Apr 17.

29. Hu Y, Sun J, Dai, Z., et al. T Prevalence and severity of corona virus disease 2019 (COVID-19): A systematic review and meta-analysis. J.Clin.Viro. 2020 Apr 14;1217.

30. Li Y, Xia L. T Coronavirus Disease 2019 (COVID-19): Role of Chest CT in Diagnosis and Management. AJR Am J Roentgenol. 2020 Mar 04; 10.2214/AJR.20.22954.

31. Chung M, Bernheim A, Mei, X., et al. T CT Imaging Features of 2019 Novel Coronavirus (2019-nCoV). RADIOLOGY. 2020 2020-01-01:200230.

32. Huang C, Wang Y, Li, X., et al. T Clinical features of patients infected with 2019 novel coronavirus in Wuhan, China. Lancet (London, England) 2020; 395(10223): 497–506.

33. Wynants L, Van Calster B, Bonten MMJ, et al. T Prediction models for diagnosis and prognosis of covid-19 infection: systematic review and critical appraisal. BMJ. 2020 04 07;369; 10.1136/bmj.m1328.

34. Zirui M, Minjin W et al. T Development and utilization of an intelligent application for aiding COVID-19 diagnosis. BMJ. 2020.03.18; https://doi.org/10.1101/2020.03.18.20035816

35. Cong F, MD, Zhi H et al. T A Novel Triage Tool of Artificial Intelligence Assisted Diagnosis Aid System for Suspected COVID-19 pneumonia In Fever Clinics. BMJ. 2020 03 19; https://doi.org/10.1101/2020.03.19.20039099

36. Fantoni M, Murri R, Scoppettuolo G, et al. T Resource-saving advice from an infectious diseases specialist team in a large university hospital: an exportable model?. Future Microbiology, 2015, 10(1):15–20.

